# Accelerated vs. Daily TMS: Resting-State Functional Connectivity in a Within-Subjects Crossover Trial

**DOI:** 10.64898/2026.11.17.26368336

**Authors:** Jillian G. Connolly, Sophia H. Blyth, Libby C. Tunison, Maxwell J. Roeske, Simon Vandekar, Baxter P. Rogers, Roscoe O. Brady, Heather Burrell Ward

**Affiliations:** Department of Psychiatry and Behavioral Sciences, Vanderbilt University Medical Center, Nashville, TN; Department of Biostatistics, Vanderbilt University Medical Center, Nashville, TN; Department of Psychiatry, Beth Israel Deaconess Medical Center and Harvard Medical School, Boston, MA and McLean Hospital, Belmont, MA

## Abstract

**Importance:** Accelerated protocols have emerged over the past two decades as a promising method of transcranial magnetic stimulation (TMS) administration that allows for shorter treatment duration and faster time to response. Although recent research has suggested accelerated TMS may be noninferior to daily TMS on clinical outcome measures, no studies have directly compared the neural effects of daily and accelerated TMS within the same participant group.

**Objective:** Compare the neural effects of daily vs accelerated TMS applied to a personalized default mode network (DMN) target on bilateral parietal DMN connectivity in a transdiagnostic sample (n=36) before and after TMS.

**Design:** Within-subjects crossover design.

**Setting:** Vanderbilt University Medical Center, Nashville, TN.

**Participants:** 36 individuals with a range of psychiatric conditions were enrolled (e.g., psychosis, depression, anxiety). Data from 34 participants are included in analysis.

**Intervention:** 5 daily sessions of DMN-targeted continuous theta burst stimulation (cTBS) with pre-/post neuroimaging. After a washout period (mean =379.26 days), these same participants returned for 5 accelerated sessions of DMN-targeted cTBS in one day with pre-/post neuroimaging.

**Main Outcome and Measure:** To compare the neural effects of accelerated versus traditional daily TMS schedules, the primary outcome measure was change in resting-state bilateral parietal DMN connectivity between pre-TMS and post-TMS.

**Results:** Across all participants and both treatment schedules, DMN-targeted cTBS reduced bilateral parietal DMN connectivity (t(90.16)=-2.07, p=0.0416). TMS schedule did not did not have a statistically significant effect in DMN connectivity change(t(31.27) = −0.44, p = 0.67) . Psychosis diagnosis did not affect bilateral parietal DMN connectivity change (t(28.34)=-0.53, p=0.601). However, older age was associated with greater reduction in bilateral parietal DMN connectivity across the entire sample (t(26.91)=-2.21, p=0.036).

**Conclusion and Relevance:** This study suggests accelerated and daily TMS protocols may have comparable neural effects across psychiatric diagnoses. Understanding the effects of TMS schedule is critical for protocol design and clinical TMS application.

**Trial Registration:** NCT07155096 (https://clinicaltrials.gov/study/NCT07155096), NCT06389266 (https://clinicaltrials.gov/study/NCT06389266)

## Introduction

Over the past forty years, transcranial magnetic stimulation (TMS) has emerged as a revolutionary treatment option for a range of psychiatric disorders, notably treatment refractory major depressive disorder (MDD) and obsessive-compulsive disorder (OCD). Up to 58% of patients who receive TMS treatment for MDD show clinical improvement (1), and TMS for OCD consistently elicits higher response rates than sham stimulation (2). Thus, TMS provides a valuable treatment alternative for patients who have had limited success with prior medications and psychotherapies.

Despite the established clinical benefits of TMS, traditional protocols require patients to attend daily sessions of TMS over the course of many weeks, placing considerable logistic and financial demands on patients and families (3,4). In 2008, the first Food and Drug Administration (FDA)- cleared protocol for MDD required patients to attend one session of TMS per day, five days per week, for 4-6 weeks (5). Over a decade later, TMS gained FDA clearance for the treatment of OCD with a 6-week treatment course of Monday-Friday daily TMS (6). Amidst these FDA approvals, the need for shorter treatment durations resulted in several clinical trials investigating “accelerated” protocols, which have transformed the delivery of TMS (7,8). Rather than administering TMS daily for weeks at a time, accelerated protocols administer multiple TMS sessions each day, which allows for shorter treatment courses and faster time to response.

Although accelerated TMS protocols date back to 2001 (9), they became increasingly common in the mid 2010s in novel trials testing TMS applied to the dorsolateral prefrontal cortex (DLPFC) for individuals with treatment-resistant depression (10,11).

The first randomized, head-to-head comparison of accelerated versus standard TMS was published in 2018 by Fitzgerald et al. (12). In this comparison, outpatients with MDD were randomized to accelerated TMS (3 sessions/day tapered over 3 weeks, 63,000 total pulses) or standard TMS (once daily x 4 weeks, 63,000 total pulses). Authors observed no significant differences in response rates, remission rates, or depression score reduction between groups, indicating that accelerated TMS may be clinically comparable to traditional daily schedules.

The publication of the Stanford Accelerated Intelligent Neuromodulation Accelerated TMS (SNT) protocol further advanced accelerated TMS, with 10 intermittent theta burst stimulation (iTBS) sessions per day delivered over 5 consecutive days for the treatment of MDD (7,8). This finding demonstrated the tolerability and effectiveness of TMS at high dosages over shorter periods of time compared to standard, daily TMS.

To further examine the clinical outcomes of accelerated versus daily TMS, Hanlon et al performed a multisite, randomized, blinded non-inferiority trial (n=104) comparing accelerated iTBS (5 sessions/day x 6 days with H1 coil, 1800 pulses/session) to standard once/daily 18 Hz TMS (1980 pulses/day x 4 weeks with H1 coil) (13). They observed accelerated iTBS was non- inferior to daily 18 Hz TMS based on change in depression scores and Week 6 response and remission rates. Moreover, accelerated treatment achieved a faster time to remission (21 vs. 28 days, p=.03). Analysis of patient-reported outcome measures from the same trial also found comparable improvements in depression, anxiety, and quality of life (14). Most recently, Apostol and colleagues published a retrospective comparison of accelerated TMS (5 sessions/day x 5 days, n=40) compared to standard once-daily TMS for at least 30 days (n=135) and found no differences in PHQ-9 scores between the two groups (15). Importantly, the accelerated iTBS group had a shorter time to response (11 vs. 16 days, p=.04), demonstrating further clinical benefit of accelerated TMS.

In summary, a growing body of evidence suggests that accelerated TMS administration is noninferior to standard daily administration for clinical outcome measures. However, a major gap in the field exists: no studies have directly compared the neural effects of accelerated and standard daily TMS protocols in the same trial, a gap highlighted by experts in the field (16). Although accelerated TMS protocols have shown transdiagnostic efficacy across psychiatric and substance use disorders (17,18), the extant literature has focused on TMS applied to the left DLPFC for depression (19). To be explicit, no studies have compared accelerated vs. daily TMS protocols in psychiatric disorders outside of depression or to alternative TMS targets. Therefore, we compared the effects of accelerated versus daily TMS applied to a personalized, parietal TMS target in a transdiagnostic sample of individuals with a variety of psychiatric disorders and substance use (n=36, Figure 1) in a secondary analysis of data from two clinical trials.

**Figure 1.**
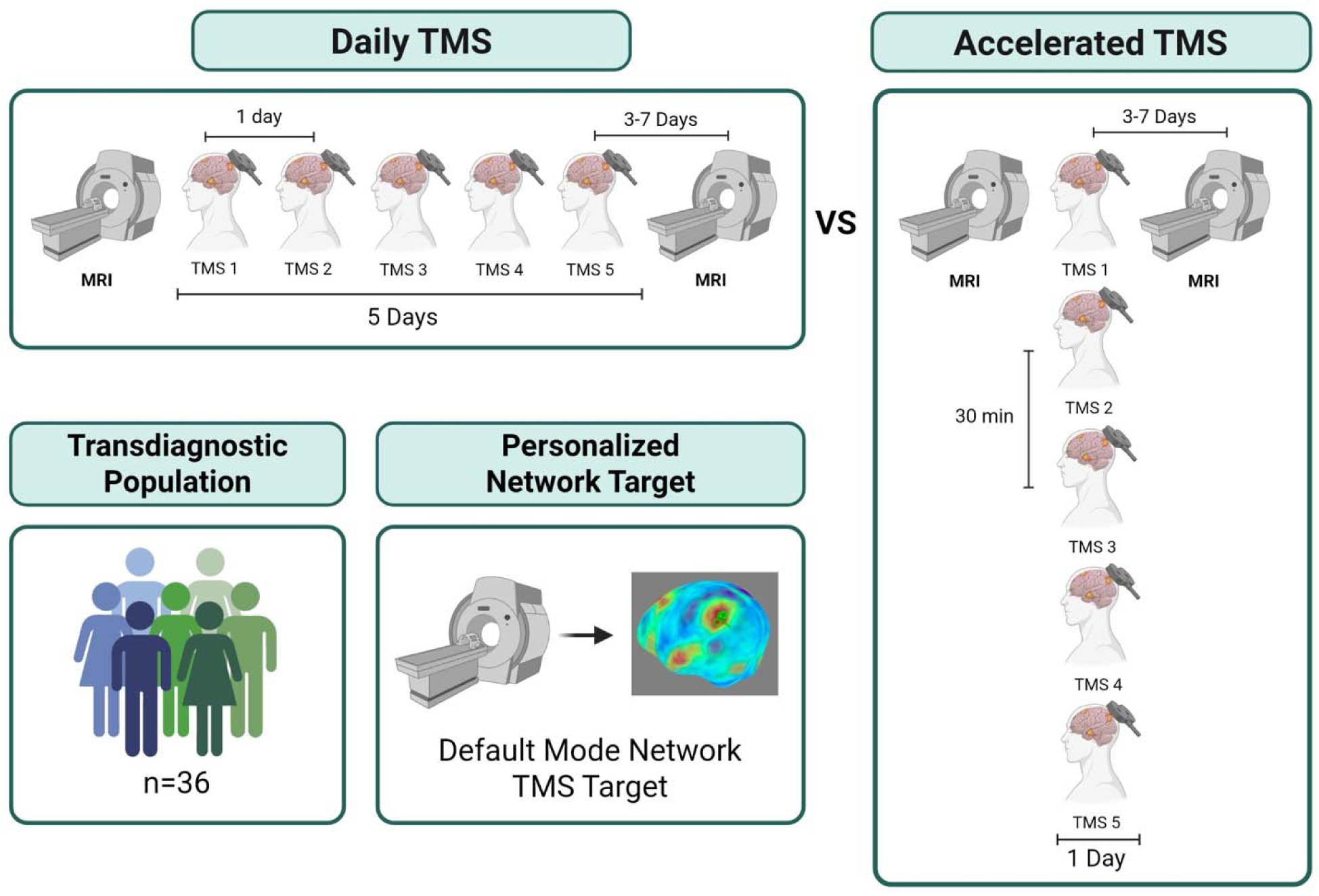
Daily vs Accelerated TMS Study Design. In this crossover trial, 36 individuals received five daily sessions of DMN-Targeted cTBS and five accelerated sessions of DMN- targeted cTBS with neuroimaging before and after each TMS intervention. Both daily and accelerated DMN-Targeted cTBS (600 pulses, 100% AMT) were applied to an individualized left parietal DMN target (see *Individualized DMN Target* in Supplement). Accelerated TMS was administered in one-day with a 30-minute inter-session interval. Daily and accelerated TMS occurred at least 3 months apart to avoid carryover effects (mean =379.26 days, range: min = 121 days, max = 972 days). Details in supplement.

Although there are many ways to compare the neural effects of accelerated and daily TMS protocols, we selected a neural outcome (bilateral parietal connectivity) that we have shown in multiple prior studies is modulated by single and multiple sessions of parietal TMS and is associated with behavior change (20,21).

This analysis fills a critical gap in the TMS literature and clinical practice. If the neural effects of accelerated and daily TMS are comparable, this would provide the first neurobiological evidence suggesting it is the cumulative dose of TMS, rather than treatment schedule, that determines the neural effects of TMS.

## Methods

### Participants

A transdiagnostic sample of 36 individuals aged 18-65 were enrolled (Table 1, Supplemental Table 1). Participants were recruited in two diagnostic groups: 1) Psychosis and 2) Non-psychosis (NCT06389266, NCT07155096). Individuals in the psychosis group had a DSM- 5 diagnosis of schizophrenia or schizoaffective disorder, while the non-psychosis group had no lifetime history of psychosis but had other psychiatric disorders (e.g., depression, anxiety, attention deficit hyperactivity disorder, see Supplemental Table 2 for diagnoses). Participants provided written informed consent. Details in supplement.

**Table 1.** Demographics. 36 individuals aged 18-65 enrolled in this study and are included in the chart above. Participants were recruited in two groups: 1) Psychosis and 2) Non-psychosis group (NCT06389266, NCT07155096). Individuals in the psychosis group had a DSM-5 diagnosis of schizophrenia or schizoaffective disorder, while individuals in the non-psychosis control group had no lifetime history of psychosis (see Supplemental Table 2 for diagnoses). Details in supplement.

| **Variable** | **Total Sample (n=36)** |
|---|---|
| Age, y (SD) | 35.4 (12.1) |
| Sex, n (% Female) | 11 (30.6%) |
| Black/African American | 10 (27.8%) |
| American Indian/Alaska Native | 1 (2.8%) |
| White | 21 (58.3%) |
| Native Hawaiian or Other Pacific Islander | 0 (0%) |
| Asian | 0 (0%) |
| Multi Racial | 3 (8.3%) |
| Other, not specified | 1 (2.8%) |
| Unknown/Missing | 0 (0%) |
| AMT | 100% |
| cTBS Intensity, Daily (SD) | 43.6 (13.6) |
| cTBS Intensity, Accelerated (SD) | 46.3 (15.2) |
| BPRS, Daily (SD) | 34.7 (8) |
| BPRS, Accelerated (SD) | 33.6 (6.6) |

### MRI Acquisition & Processing

Imaging data were collected on 3.0-T Philips Intera Achieva MRI scanner (Philips Healthcare, Andover, MA). Participants completed MRIs the week before and after TMS. Post-TMS scans were scheduled within 8 days of TMS (daily mean 4.39 (SD 1.61) days; accelerated mean 4.91 (SD 1.38) days, p=0.153). Briefly, 1-mm^3^ T1-weighted anatomical scans and multiple 10-minute functional runs were acquired (TR 2000ms, TE 28.0ms, flip angle 90 degrees, field of view = 240mm, 38 slices, 3-mm^3^ voxels, anterior to posterior phase-encoded). Scans were processed using the same procedure described above. Details in supplement. After quality control, there were 126 pre-/post-TMS scans for 34 participants.

### TMS Protocol

In this crossover trial, individuals received five daily sessions of DMN-Targeted continuous theta burst stimulation (cTBS) and five accelerated sessions of DMN-targeted cTBS.

TMS applied using a MagPro X100 stimulator and active figure-of-8 coil (Cool B65, MagVenture, Denmark) held tangentially to the scalp with the handle at 45 degrees. TMS was applied in the standard theta-burst pattern (3 pulses at 50-Hz repeated at a rate of 5-Hz for 600 pulses at 100% AMT, details in Supplement) (22). Both daily and accelerated DMN-Targeted cTBS were applied to an individualized left parietal DMN target (see *Individualized DMN Target*). Accelerated TMS was administered in one-day with a 30-minute inter-session interval. Daily and accelerated TMS occurred at least 3 months apart to avoid carryover effects (mean =379.26 days, range: min = 121 days, max = 972 days), consistent with prior literature (23).

### Individualized DMN Targeting

To identify an individualized DMN map for TMS targeting, a standard DMN template (24) was warped into native space and applied to the participant’s pre- TMS scan. In each participant, the resultant connectivity maps yielded a correlation cluster in the left posterior inferior parietal lobule (IPL). A target was placed in the averaged center of the left posterior IPL correlation cluster (formed from the overlay of the left posterior IPL clusters derived from connectivity maps) on the cortical surface using Brainsight neuronavigation software (Rogue Research, Inc.). This individualized left parietal DMN region served as the TMS target for both daily and accelerated TMS.

### Region-of-Interest (ROI) Neuroimaging Analysis

To compare neural effects of accelerated and daily TMS protocols, we selected a neural outcome (bilateral parietal connectivity) that we have shown in multiple prior studies is modulated by single and multiple sessions of personalized, parietal TMS and is associated with behavior change (20,21). To calculate bilateral parietal connectivity, we extracted time courses from a 6mm sphere placed at our left lateral parietal DMN TMS target (x = -46, y = -66, z = +30) (Raichle 2011) and the right parietal DMN region we have previously linked to TMS-induced neural change and behavior change (25) The BOLD signal time courses from ROIs were correlated with each other and z-transformed to generate ROI-to-ROI connectivity values for analysis. See Supplemental Methods for Details.

### Statistical Analysis

To compare DMN connectivity before and after TMS, linear mixed models were used to predict bilateral parietal DMN connectivity based on time, group, TMS schedule (accelerated vs daily), and age with a random effect for subject. All analyses were conducted in RStudio version 2025.09.2. See Supplemental Tables 3-7 for details.

## Results

Thirty-four participants completed both daily and accelerated DMN-targeted cTBS and were included in analysis. Nineteen participants were diagnosed with psychotic disorders, and fifteen were diagnosed with non-psychotic psychiatric disorders (Supplemental Table 2). No serious adverse events were reported (see Supplemental Tables 8-9 for minor adverse events).

### DMN-Targeted cTBS Reduces Bilateral Parietal DMN Connectivity

In a model predicting bilateral parietal DMN connectivity based on visit (pre- or post-TMS) and a random effect for subject, bilateral parietal DMN connectivity was significantly reduced following DMN-targeted cTBS administration across all participants and both treatment schedules (t(90.16)=-2.07, p=0.0416) (Figure 2A, Supplemental Table 3). In a separate linear mixed effects model examining bilateral parietal DMN connectivity change based on group (psychosis vs non-psychosis), there was no effect of group (t(28.34)=-0.53, p=0.601) on DMN connectivity change (Figure 2B, Supplemental Table 4).

**Figure 2.**
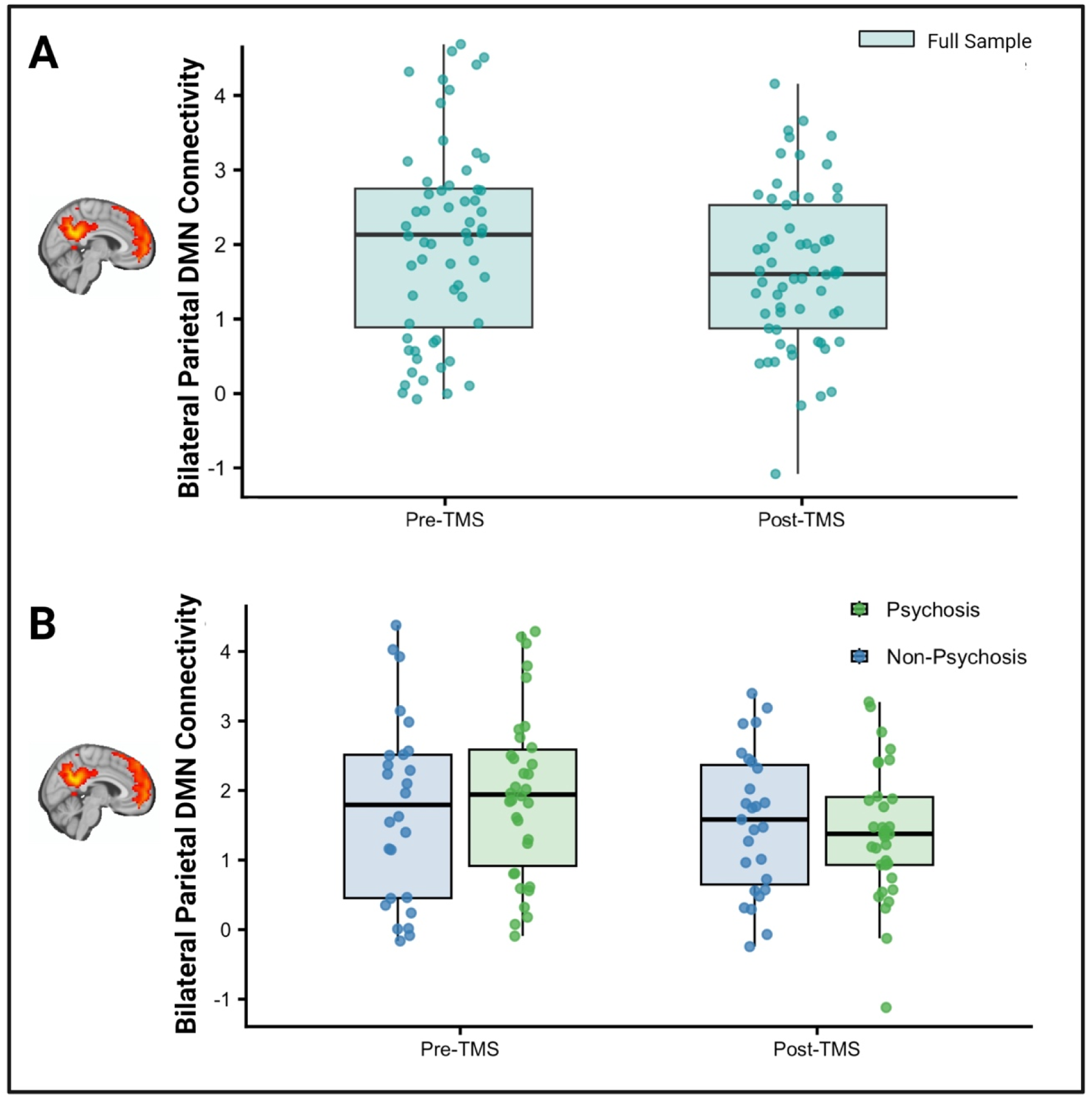
DMN-Targeted cTBS significantly reduces bilateral parietal DMN connectivity. A.) In a model predicting bilateral parietal DMN connectivity change based on visit (pre- or post-TMS) and including a random effect for subject, bilateral parietal DMN connectivity was significantly reduced following DMN-targeted cTBS administration across all participants and both treatments schedules (t(90.16)=-2.07, p=0.0416). B.) In a separate linear mixed effects model examining bilateral parietal DMN connectivity change based on group (psychosis or non- psychosis group), no significant differences in bilateral parietal DMN connectivity change were observed between the psychosis group and the non-psychosis groups (t(28.34)=-0.53, p=0.601).

### Neural Effects of TMS Do Not Differ by TMS Administration Schedule

To test the effects of TMS schedule (daily vs accelerated) on bilateral parietal DMN connectivity change, a linear mixed effects model was used which modeled bilateral parietal DMN connectivity change based on the pre-TMS bilateral parietal DMN connectivity and TMS schedule while including a random effect for subject. TMS schedule (t(31.27)= -0.44, p=0.67, Cohen’s d = 0.12, 95% CI [-0.45, 0.69]) was not a significant predictor of bilateral parietal DMN connectivity change, indicating that no significant differences in connectivity change were observed between accelerated and daily TMS schedules (Figure 3A). Only pre-TMS bilateral parietal DMN connectivity predicted post-TMS bilateral parietal DMN connectivity change (t(55.94)=-7.82, p<0.001) (Supplemental Table 5).

**Figure 3.**
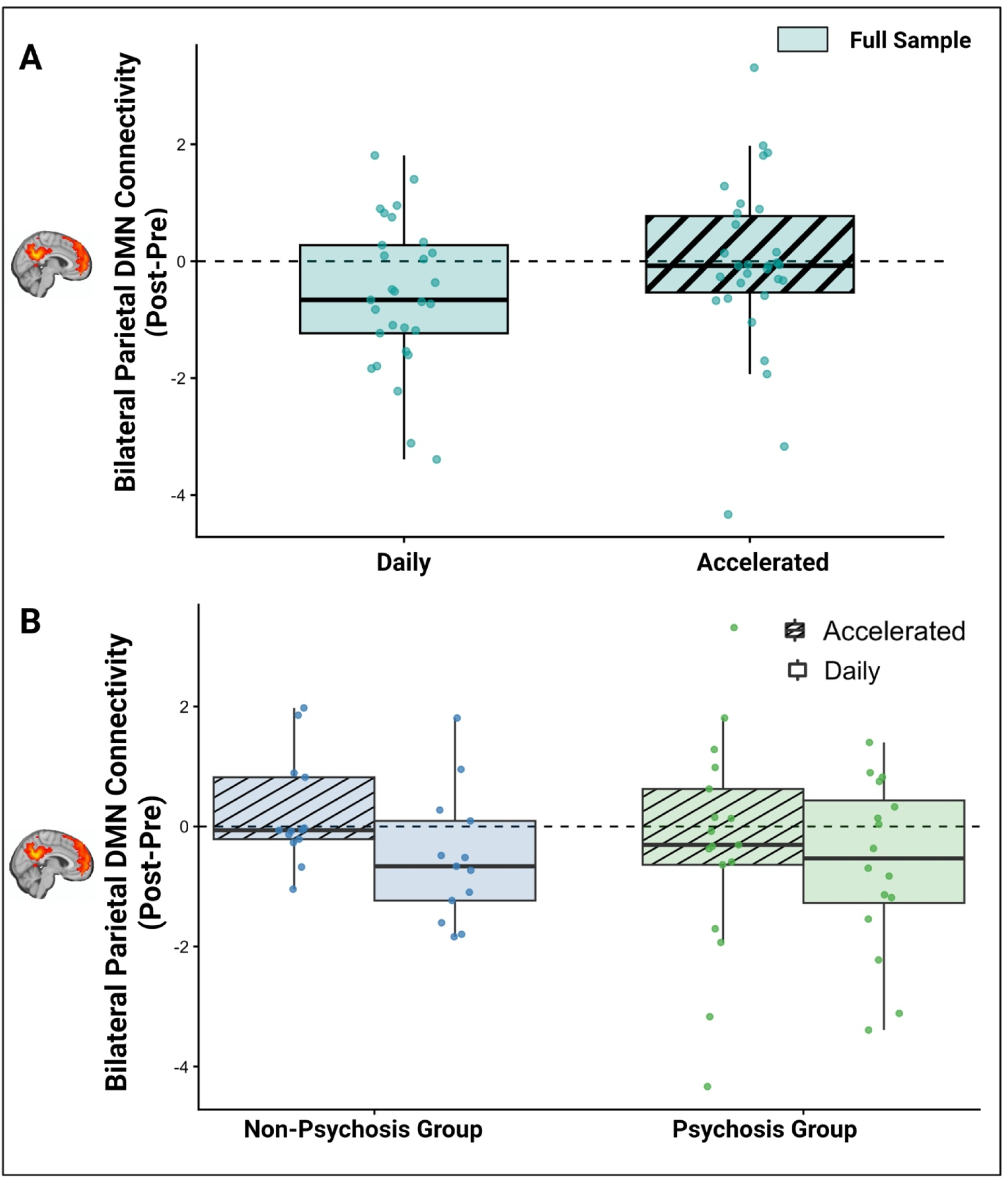
Changes in bilateral parietal DMN connectivity following cTBS in a transdiagnostic sample. TMS schedule (accelerated TMS vs daily TMS) was not associated with significant differences in bilateral parietal DMN connectivity change between the psychosis group and the non-psychosis group: In a model testing bilateral parietal DMN connectivity change based on pre-TMS bilateral parietal DMN connectivity and the TMS schedule*group interaction, TMS schedule (t(28.10)=-1.09, p=0.284), group (t(44.52)=-1.01, p=0.318), and the TMS schedule*group (t(26.73)=1.07, p=0.292), interaction were not significant predictors of connectivity change; only pre-TMS bilateral parietal DMN connectivity predicted post-TMS DMN connectivity change (t(53.97)=-7.75, p<0.001).

Diagnosis did not interact with TMS schedule (accelerated TMS vs daily TMS) to affect bilateral parietal DMN connectivity change. In a model predicting bilateral parietal DMN connectivity change based on pre-TMS bilateral parietal DMN connectivity and the TMS schedule*group interaction, TMS schedule (t(28.10)=-1.09, p=0.284), group (t(44.52)=-1.01, p=0.318), and the TMS schedule*group (t(26.73)=1.07, p=0.292), interaction were not significant predictors of connectivity change; only pre-TMS bilateral parietal DMN connectivity predicted post-TMS bilateral parietal DMN connectivity change (t(53.97)=-7.75, p<0.001) (Figure 3B, Supplemental Table 6).

### Neural Effects of TMS May Differ by Age

In an exploratory analysis to investigate the effects of age on neural response to TMS, bilateral parietal DMN connectivity change was modeled based on pre-TMS bilateral parietal DMN connectivity, TMS schedule, group, and age while including a random effect for subject. Age (t(26.91)=-2.21, p=0.036) and pre-TMS bilateral parietal DMN connectivity (t(53.28)=-7.97, p<0.001) were both significant predictors of bilateral parietal DMN connectivity change following TMS. Older age was associated with greater reductions in bilateral parietal DMN connectivity across all participants and both treatment schedules (Figure 4A & 4B, Supplemental Table 7).

**Figure 4.**
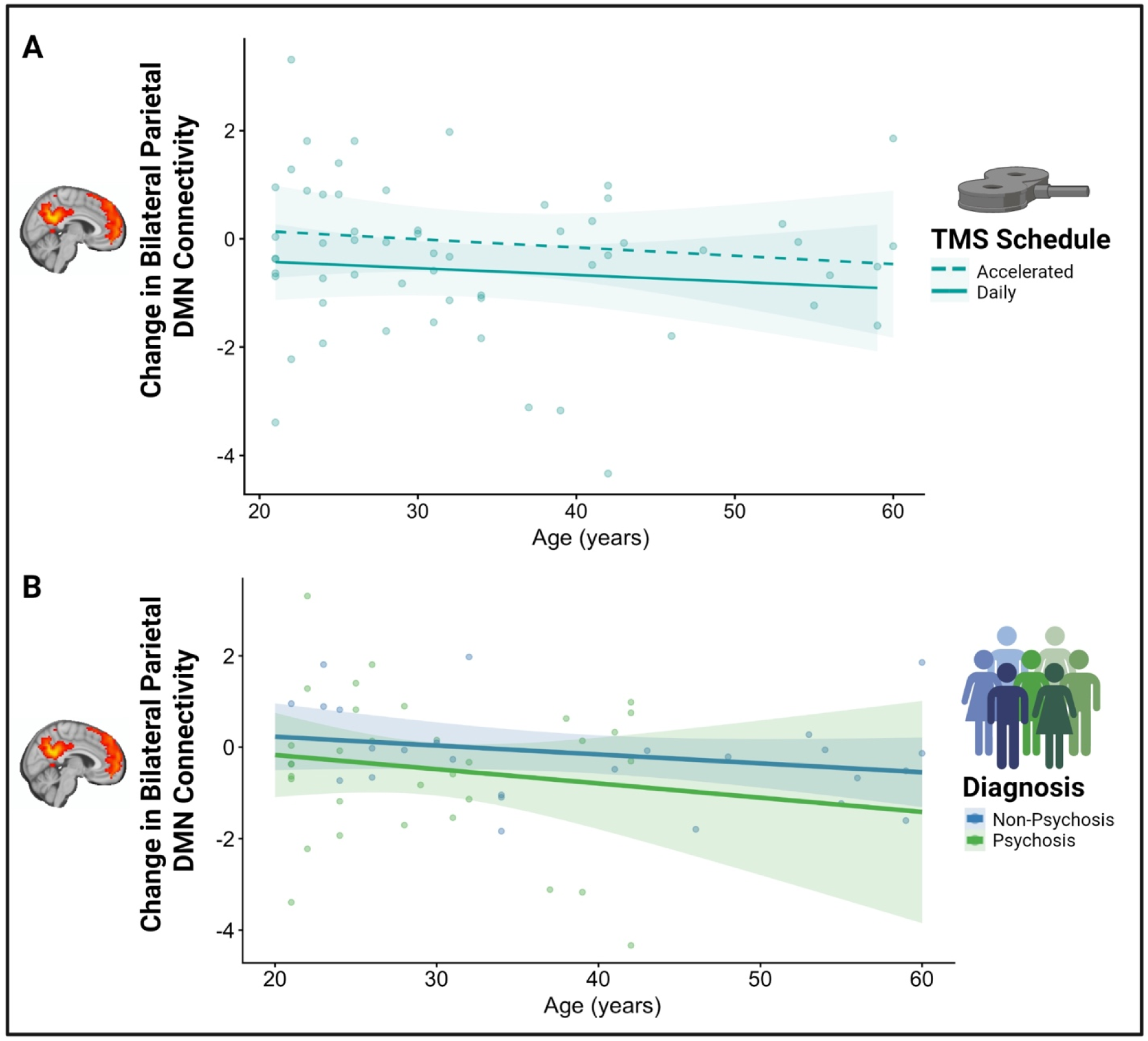
Older age is associated with greater reductions in bilateral parietal DMN connectivity following DMN-targeted cTBS regardless of treatment schedule or diagnostic group. Bilateral Parietal DMN connectivity change was modeled based on pre-TMS bilateral parietal DMN connectivity, TMS schedule, group, and age while including a random effect for subject. Age (t(26.91)=-2.21, p=0.036) and pre-TMS bilateral parietal DMN connectivity (t(53.28)=-7.97, p<0.001) were both significant predictors of bilateral parietal DMN connectivity change following TMS. Older age was associated with greater reductions in bilateral parietal DMN connectivity across all participants and both treatment schedules.

## Discussion

Accelerated TMS has been heralded since the publication of Stanford Neuromodulation Therapy in 2020 (7,8). Growing evidence suggests that accelerated TMS protocols are noninferior to daily administration for clinical outcomes. However, no studies have compared the neural effects of accelerated versus daily TMS administration.

This is the first study comparing neural effects of accelerated versus daily TMS administration. We used a within-subjects crossover design in a transdiagnostic psychiatric sample and observed that TMS schedule was not associated with connectivity change, suggesting that accelerated and daily TMS may have similar neural effects.

Our findings are consistent with the growing body of literature suggesting that accelerated and daily TMS protocols may have comparable effects. Several studies have shown that accelerated and daily TMS have comparable anti-depressant effects. Our study now suggests that the comparable clinical effects may be due to comparable neural effects of accelerated and daily TMS schedules. This also supports a broader principle in TMS treatment: the total dose of stimulation determines response, rather than the schedule on which it is administered. If this principle is true, it would follow that spaced TMS (i.e., less than daily) would also have the same effects as daily TMS. This was tested by Galletly et al, who compared spaced TMS (3 days/week x 6 weeks) to daily TMS (5 days/week x 4 weeks) for major depression (26). At the end of treatment, they observed no differences in antidepressant response between groups, suggesting response was determined by was total number of treatments, rather than treatment schedule.

However, they found that at 4 weeks, the daily group (which had already received 20 treatments compared to the spaced group, which had only received only 12) had a faster response. This suggests that response to TMS may be predicted by number of sessions rather than administration schedule, whether daily, accelerated, or spaced.

Across all diagnoses and both treatment schedules, older age was associated with greater reductions in bilateral parietal DMN connectivity following TMS. While exploratory, this finding is consistent with previous literature which associates higher age with greater improvement in depression severity following TMS (27–29). Given evidence for age related changes in DMN connectivity (30), it is possible that TMS treatment response may differ between older and younger adults. However, future studies are needed to directly examine the role of age on treatment efficacy and to identify the neural mechanism underlying these changes.

This study has several major strengths. The primary strength of this study was its within-subjects crossover design that compares the neural effects of accelerated versus daily TMS administration within the same participant, thereby minimizing confounding from between-subjects factors.

Retention of highly symptomatic individuals across two interventions with repeat neuroimaging and an extended washout period (mean =379.26 days) was another major strength. Each participant received four MRI scans, providing dense within-subject sampling. Furthermore, individualized TMS targets were created from each participant’s resting state functional MRI scans, ensuring that stimulation was personalized to each participant’s DMN. This approach represents a unique methodological strength, as recent evidence suggests that personalized, connectivity-based targeting may lead to greater clinical benefit than scalp landmark-based targeting (17). Another strength of this study was the use of a transdiagnostic psychiatric sample that included individuals with psychotic, mood, anxiety, and substance use disorders, an approach used in other TMS trials (31–35). Given evidence that accelerated TMS has transdiagnostic effects (18), our analysis provides evidence that the neural effects of accelerated and daily TMS administration do not differ regardless of diagnosis. However, our sample was not powered to investigate diagnostic differences, so this should be explored in future studies.

Our study also has several limitations. First, we investigated the effects of five sessions of TMS, and it is possible that neural effects between accelerated and daily TMS may differ after more sessions. However, testing a smaller number of sessions, where the duration of TMS effect is shorter allowed us to conduct this unique crossover design trial. It is important to note that this study was conducted using a fixed-order design where each participant received daily TMS before accelerated TMS. Although we cannot fully exclude the possibility that treatment order contributed to the results, the three-month minimum washout period (mean 379.26 days) separating the two arms well exceeds the timeframe that TMS-induced functional connectivity changes are expected to persist. Previous work has demonstrated that neural effects are expected to normalize on a scale of hours to days following single sessions of cTBS (36). There is little evidence to suggest that 5 single sessions of TMS would persist beyond 3 months (37). However, to fully address this limitation, future studies using a counterbalanced crossover design are needed. An additional limitation is that this study was not powered as a formal noninferiority trial. Although no significant differences in neural outcomes were observed between accelerated and daily TMS, the relatively small sample size resulted in wide confidence intervals that do not exclude clinically meaningful differences in connectivity change. Larger, adequately powered noninferiority studies are needed to determine whether accelerated and daily TMS produce formally comparable neural outcomes within prespecified noninferiority margins.

Another limitation of our findings was the use of a personalized, network-based target. While our finding that accelerated and daily TMS may have similar neural effects is consistent with studies investigating the clinical outcomes from DLPFC-targeted TMS, it is possible that our findings are specific to TMS applied to the parietal cortex. Future studies should compare accelerated versus daily TMS administration using different TMS targets. Finally, while our transdiagnostic sample enhances the generalizability of our findings, it is also a limitation. It is possible that there are differences in accelerated versus daily TMS administration by diagnostic groups. It is also possible that differences in neural response differ person-to-person rather than by diagnostic group.

Due to the time and transportation barriers that many patients face with tedious, daily TMS treatment (3,4), accelerated TMS protocols offer the opportunity for increased convenience and access to care by reducing the number of treatment visits. In addition to shortening the treatment course, accelerated TMS protocols have been shown to expedite treatment response compared to traditional daily TMS protocols (12). Given the clear practical and clinical benefits of accelerated TMS, there has been increased interest in the therapeutic benefit of daily TMS schedules compared to accelerated TMS schedules. However, major questions remain about administration parameters for accelerated TMS, including the optimal number of pulses per session, number of sessions per day, and inter-session interval that should be explored in future studies (17,19).

In conclusion, this is the first study to compare the neural effects of accelerated and daily TMS administration. The present study provides preliminary evidence suggesting TMS treatment schedule may not affect neural response to TMS across a range of psychiatric conditions, which may inform TMS protocol development and clinical implementation.

## Data Availability

Data are available upon reasonable request for an approved purpose with a signed data use agreement.

## Acknowledgments

This work was supported by funding from the Charlotte and Donald Test Fund, the Vanderbilt Kennedy Center, and the National Institutes of Health grant K23DA059690 to Dr. Ward.

