## Supplement for "Accelerated vs. Daily TMS: Resting-State Functional Connectivity in a Within-Subjects Crossover Trial"

**Supplemental Material**

**Supplemental Methods**

*Participants:* 36 individuals were enrolled in this transdiagnostic sample. 34 individuals completed TMS. These participants were divided into either a psychosis (n=19) or non-psychosis group (n=15) and all used nicotine. In the psychosis group, diagnosis of schizophrenia or schizoaffective disorder was confirmed by DSM-5 SCID interview (1) and clinical information obtained from outpatient psychiatric providers. In the non-psychosis group, diagnoses were obtained from clinical information from outpatient psychiatric providers (Supplemental Table 2). For one month prior to enrollment, individuals received outpatient care, with no hospitalizations or changes to their psychiatric medication regimens. Individuals were excluded if they had DSM-5 intellectual disability, substance use disorder (other than nicotine) in the past 3 months, a progressive or genetic neurologic disorder, history of significant head trauma, history of seizures or neurosurgical procedures, implanted devices, gross organic pathology on neuroimaging, contraindications to MRI or TMS, or current pregnancy. All participants provided written informed consent in accordance with the Vanderbilt University Medical Center Institutional Review Board.

*MRI Acquisition and Data Processing:* Imaging data were collected on 3.0-T Philips Intera Achieva MRI scanner (Philips Healthcare, Andover, MA). Participants completed MRI scans before and after the TMS intervention. Post-TMS scans were scheduled within 8 days of TMS (daily mean 4.39 (SD 1.61) days; accelerated mean 4.91 (SD 1.38) days, p=0.153). Briefly, 1-mm^3^ T1-weighted anatomical scans and multiple 10-minute functional runs were acquired (TR 2000ms, TE 28.0ms, flip angle 90 degrees, field of view = 240mm, 38 slices, 3-mm^3^ voxels, anterior to posterior phase-encoded). Anatomical images were segmented into GM, WM, and CSF with the Computational Anatomy Toolbox 12 (CAT12, version 12.5; [http://www.neuro.uni-jena.de/cat/](https://nam12.safelinks.protection.outlook.com/?url=http%3A%2F%2Fwww.neuro.uni-jena.de%2Fcat%2F&data=05%7C01%7Cheather.b.ward%40vumc.org%7C53146c81d5534edf7b5208db2189eebc%7Cef57503014244ed8b83c12c533d879ab%7C0%7C0%7C638140648518151829%7CUnknown%7CTWFpbGZsb3d8eyJWIjoiMC4wLjAwMDAiLCJQIjoiV2luMzIiLCJBTiI6Ik1haWwiLCJXVCI6Mn0%3D%7C3000%7C%7C%7C&sdata=Fsce8FRawoYi0jL0FjzH%2BUsdG8P5cAj4Wzyuk79pHuo%3D&reserved=0)). Resting-state scans were preprocessed in SPM12 (“SPM–Statistical Parametric Mapping,” <http://www.fil.ion.ucl.ac.uk/spm>)) and (1) realigned to a mean scan, (2) coregistered with the native space structural scan, then (3) underwent resting-state denoising procedures: bandpass filter (0.01–0.1 Hz), regression of CSF, WM, and mean GM signal, regression of 12 motion parameters (6 translation and rotation parameters and their first derivative, [https://github.com/baxpr/connprep/tree/38b1b04b](https://nam12.safelinks.protection.outlook.com/?url=https%3A%2F%2Fgithub.com%2Fbaxpr%2Fconnprep%2Ftree%2F38b1b04b&data=05%7C02%7Cheather.b.ward%40vumc.org%7C30709cbd856343dda76e08df35382ddc%7Cef57503014244ed8b83c12c533d879ab%7C0%7C0%7C639288187539288696%7CUnknown%7CTWFpbGZsb3d8eyJFbXB0eU1hcGkiOnRydWUsIlYiOiIwLjAuMDAwMCIsIlAiOiJXaW4zMiIsIkFOIjoiTWFpbCIsIldUIjoyfQ%3D%3D%7C0%7C%7C%7C&sdata=cIEmgRdgcbr6p6aDylA4fmd5n%2BVvdRwYZ%2FNNGDQ%2FYuk%3D&reserved=0)). All resting-state scans went through a quality assurance procedure that included calculating FD and tSNR. Scans with mean FD > 0.6 or tSNR lower than the 5th percentile of the sample distribution were excluded from analysis.

*Region-of-Interest (ROI) Neuroimaging Analysis:* To compare neural effects of accelerated and daily TMS protocols, we selected a neural outcome (bilateral parietal connectivity) that we have shown in multiple prior studies is modulated by single and multiple sessions of personalized, parietal TMS and is associated with behavior change (20,21). To calculate bilateral parietal connectivity, we extracted time courses from a 6mm sphere placed at our left lateral parietal DMN TMS target (x = -46, y = -66, z = +30) (Raichle 2011) and the right parietal DMN region we have previously linked to TMS-induced neural change and behavior change (25) The BOLD signal time courses from ROIs were correlated with each other and z-transformed to generate ROI-to-ROI connectivity values for analysis (<https://github.com/baxpr/conncalc/tree/v1.9.1>).

*TMS Motor Threshold Determination:* Participants had motor threshold determination at prior to their first TMS session in both the daily and accelerated interventions. Single pulse and repetitive stimulation was performed with a MagPro X-100 stimulator (MagVenture) equipped with a biphasic figure-of-eight coil.  To obtain an active motor threshold, single pulses were used in the following manner: Electromyographic activity (EMG) was recorded using surface electrodes attached to the skin to measure motor evoked potentials (MEP) during the motor threshold assessment. The TMS coil was placed on the scalp. Single TMS pulses were applied over the hand area of the left motor cortex and individually localized for each participant based on the optimal position for eliciting a motor evoked potential. Neuronavigation (Brainsight, Rogue Research, Inc.) was used to record the motor ‘hot-spot.’  Resting and active motor threshold (RMT; AMT) were obtained by following recommendations from the International Federation of Clinical Neurophysiology.

**Supplemental Figures and Tables**

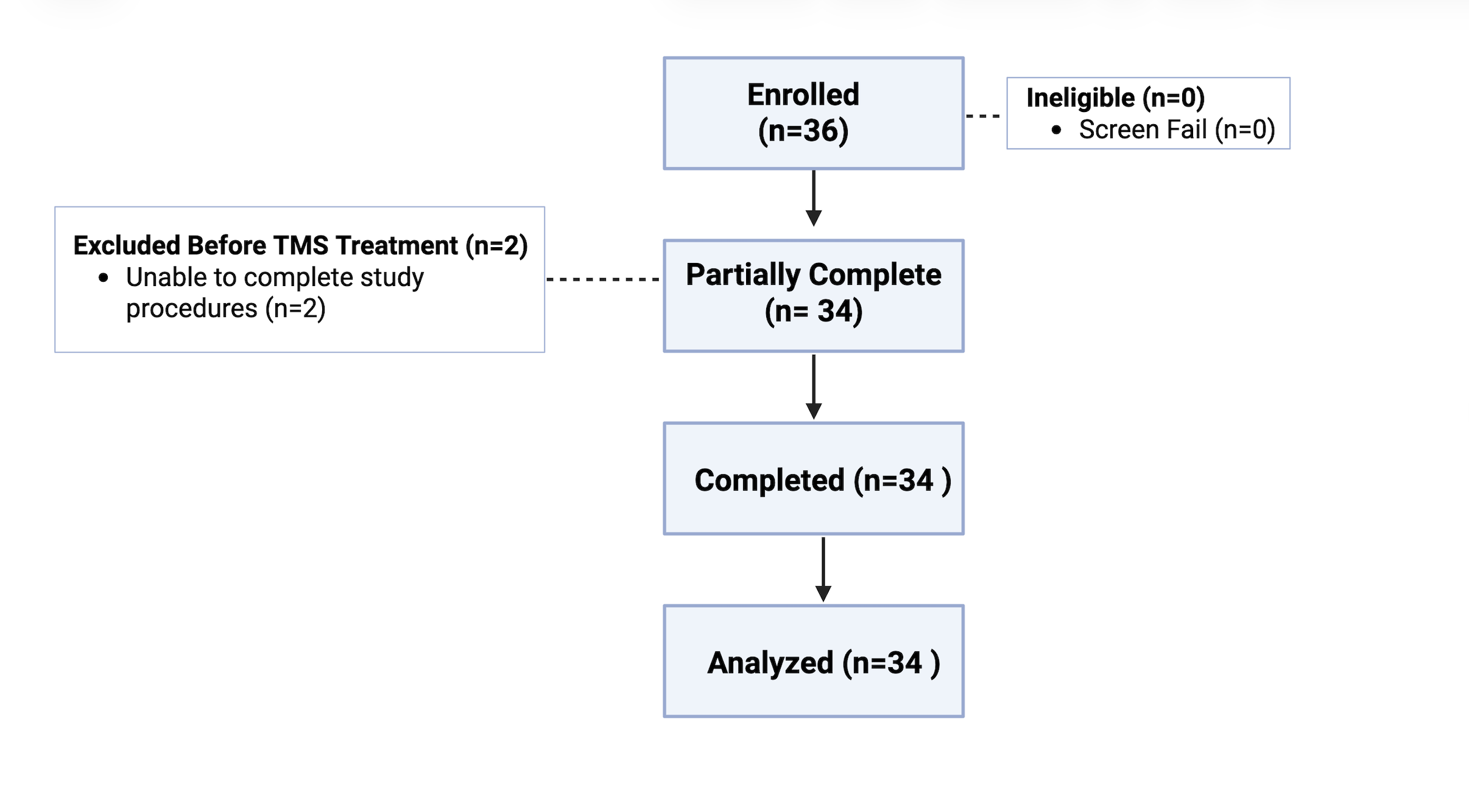

**Supplemental Figure 1.** Accelerated vs Daily DMN-Targeted cTBS CONSORT Diagram. Thirty-six participants with a range of psychiatric diagnoses aged 18-65 were enrolled in this study. Thirty-four participants completed the study procedures. Two participants were excluded before cTBS treatment because one was unable to complete an MRI and the second was unable to complete the study within the proper time constraints.

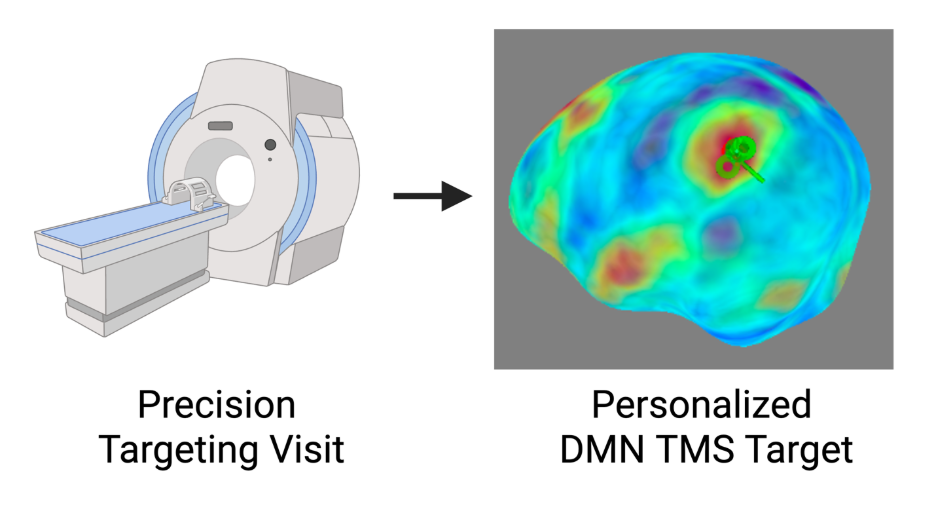

**Supplemental Figure 2. Individualized Default Mode Network TMS Target***:* The Default Mode Network (DMN) target was identified using the same methods for daily and accelerated TMS administration. For our DMN target, we selected the left lateral parietal DMN, as it is a DMN region that is readily identifiable across all individuals and has been successfully used to modulate DMN connectivity with TMS (5). To identify an individualized DMN map for TMS targeting, a standard DMN template (4) was warped into native space and applied to the participant’s baseline or pre-TMS scan (<https://github.com/baxpr/connprep/tree/38b1b04b>). The participant scan and DMN mask were then warped back into standard MNI space. In each participant, the resultant connectivity maps yielded a correlation cluster in the left posterior inferior parietal lobule. A target was then placed in the averaged center of the left posterior inferior parietal lobule correlation cluster (formed from the overlay of the left posterior inferior parietal lobule clusters derived from the connectivity maps) on the cortical surface using Brainsight neuronavigation software (Rogue Research, Inc.) An individualized TMS target was selected in the left parietal region of the DMN and used as the TMS target for all TMS sessions. Created with BioRender.com.

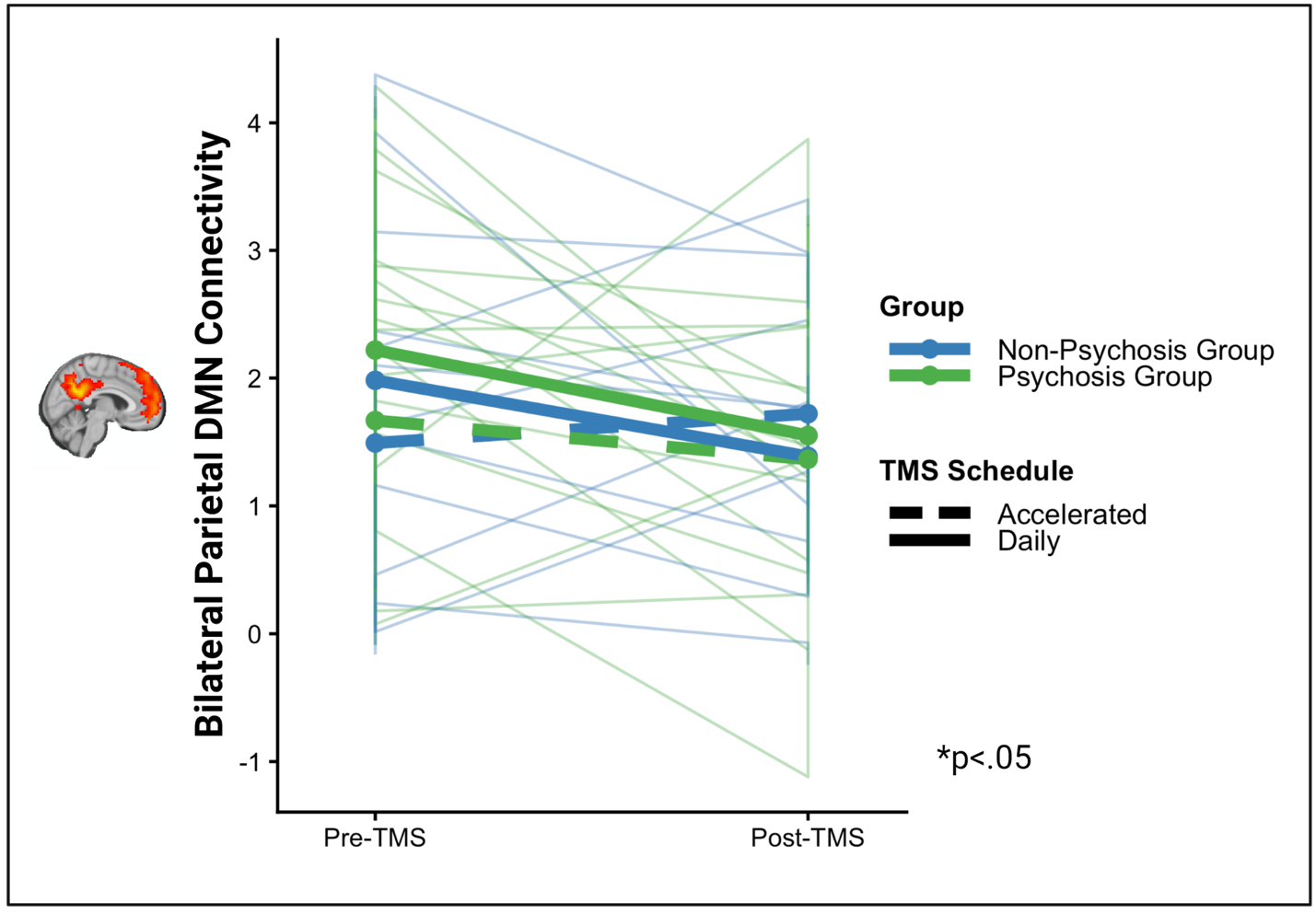
**Supplemental Figure 3. Bilateral Parietal DMN Connectivity is reduced following DMN-targeted cTBS across the transdiagnostic sample***.* Both daily and accelerated cTBS are associated with reductions in bilateral parietal DMN connectivity across psychosis and non-psychosis participants. TMS schedule (accelerated TMS vs daily TMS) was not associated with significant differences in bilateral parietal DMN connectivity change between the psychosis group and the non-psychosis group: In a model testing bilateral parietal DMN connectivity change based on pre-TMS bilateral parietal DMN connectivity and the TMS schedule*group interaction, TMS schedule (t(28.10)=-1.09, p=0.284), group (t(44.52)=-1.01, p=0.318), and the TMS schedule*group (t(26.73)=1.07, p=0.292), interaction were not significant predictors of connectivity change; only pre-TMS bilateral parietal DMN connectivity predicted post-TMS bilateral parietal DMN connectivity change (t(53.97)=-7.75, p<0.001).

| **TMS Administration by Diagnostic Group (n=36)** | | | | |
| --- | --- | --- | --- | --- |
|  | | **Psychosis**  (n=20) | **Non-Psychosis**  (n=16) | ***p*** |
| **Age, y (SD)** | | 30.8 (9.0) | 40.0 (13.7) | 0.03 |
| **Sex, n (% Female)** | | 3 (15%) | 8 (50.0%) | 0.03 |
| **Black/African American** | | 9 (45.0%) | 1 (6.2%) | 0.02 |
| **American Indian/Alaska Native** | | 0 (0%) | 1 (6.2%) | 0.441 |
| **White** | | 9 (45.0%) | 12 (75.0%) | 0.10 |
| **Native Hawaiian or Other Pacific Islander** | | 0 (0%) | 0 (0%) | 1.00 |
| **Asian** | | 0 (0%) | 0 (0%) | 1.00 |
| **Multi Racial** |  | 2 (10.%) | 1 (6.2%) | 1.00 |
| **Other, not specified** | | 0 (0%) | 1 (6.2%) | 0.44 |
| **Unknown/**  **missing** | | 0 (0%) | 0 (0%) | 1.00 |
| **BPRS Total (SD)** |  | 35.1 (6.4) | 31.7 (6.6) | 0.15 |
| **AMT** | | 44.1 (16.2) | 43.1(10) | 0.06 |
| **cTBS intensity** |  | 100% AMT | 100% AMT | - |

**Supplemental Table 1. Demographics by Diagnosis** 36 individuals were enrolled in the study, and 34 individuals aged 18-65 were included in analysis. Participants were recruited in two groups: 1) Psychosis and 2) Non-psychosis transdiagnostic group (NCT06389266). Individuals in the psychosis group had a DSM-5diagnosis of schizophrenia or schizoaffective disorder, while individuals in the non-psychosis control group had no lifetime history of psychosis (see Supplemental Table 3 for diagnoses). Participants provided written informed consent.

| **Psychosis Diagnosis (n=19)** | | |
| --- | --- | --- |
|  | Schizophrenia | 5 |
|  | Schizoaffective Disorder | 14 |
|  | Depression | 1 |
|  | Anxiety | 11 |
|  | PTSD | 2 |
|  | OCD | 1 |
|  | ADHD | 1 |
| **Non-Psychosis Diagnosis (n=15)** | | |
|  | Depression | 10 |
|  | Anxiety | 10 |
|  | ADHD | 4 |
|  | Bipolar Disorder | 1 |
|  | Borderline Personality Disorder | 1 |
|  | Adjustment Disorder | 1 |

**Supplemental Table 2. Diagnosis Breakdown of Transdiagnostic Sample (n=34).** Data was collected from 19 participants who were diagnosed with a psychotic disorder. The non-psychosis group included participants with a range of psychiatric conditions including depression (n=10), anxiety (n=10), ADHD (n=4), Bipolar Disorder (n=1), Borderline Personality Disorder (n=1), Adjustment Disorder (n=1). Participants may have met criteria for more than one diagnosis, so the diagnostic categories are not mutually exclusive.

| **Transdiagnostic Sample Diagnoses (n=34)** | | |
| --- | --- | --- |
|  | Depression | 11 |
|  | Anxiety | 21 |
|  | ADHD | 2 |
|  | Bipolar Disorder | 1 |
|  | Borderline Personality Disorder | 1 |
|  | Adjustment Disorder | 1 |
|  | PTSD | 2 |
|  | OCD | 1 |

**Supplemental Table 2. Diagnosis Breakdown of Transdiagnostic Sample (n=34).** Data was collected from 19 participants who were diagnosed with a psychotic disorder. The non-psychosis group included participants with a range of psychiatric conditions including depression (n=10), anxiety (n=10), ADHD (n=4), Bipolar Disorder (n=1), Borderline Personality Disorder (n=1), Adjustment Disorder (n=1). Participants may have met criteria for more than one diagnosis, so the diagnostic categories are not mutually exclusive.

| **Predictors** | **Estimate** | **Std. Eror** | **df** | **t-Value** | **p-value** |
| --- | --- | --- | --- | --- | --- |
| **Intercept** | 1.5016 | 0.1679 | 53.5915 | 8.944 | 3.25e-12*** |
| **Visit(preTMS =1** | -0.3493 | 0.1690 | 90.1644 | -2.067 | 0.0416 * |

**Supplemental Table 3. DMN-Targeted cTBS Reduces Bilateral Parietal** **DMN Connectivity*. Model*:** Bilateral Parietal DMN Connectivity ~ Visit + (1 | subid)

| **Predictors** | **Estimate** | **Std. Eror** | **df** | **t-value** | **p-value** |
| --- | --- | --- | --- | --- | --- |
| **Intercept** | 1.39 | 0.31 | 40.85 | 4.55 | <0.001* |
| **Pre-TMS Visit Bilateral Parietal** **DMN Connectivity** | -0.88 | 0.11 | 55.45 | -8.38 | <0.001* |
| **Group** | -0.17 | 0.33 | 28.34 | -0.53 | 0.601 |

**Supplemental Table 4. Diagnostic Group Does Not Predict Bilateral Parietal** **DMN connectivity Change Following DMN-targeted TMS. *Model:*** Bilateral Parietal DMN Connectivity Change ~ BaselineBilateral Parietal DMN connectivity + group + (1 | subid)

| **Predictors** | **Estimate** | **Std. Eror** | **df** | **t-value** | **p-value** |
| --- | --- | --- | --- | --- | --- |
| **Intercept** | 1.30 | 0.26 | 52.24 | 5.11 | **<0.001*** |
| **Pre-TMS Visit Bilateral Parietal** **DMN Connectivity** | -0.87 | 0.11 | 55.94 | -7.82 | <0.001* |
| **Schedule** | -0.09 | 0.20 | 31.27 | -0.44 | 0.67 |

**Supplemental Table 5. Neural Effects of TMS Do Not Differ by TMS Administration Schedule Following DMN-targeted TMS. *Model:*** Bilateral Parietal DMN Connectivity Change ~ Baseline Bilateral Parietal DMN Connectivity + group + (1 | subid)

| **Predictors** | **Estimate** | **Std. Eror** | **df** | **t-value** | **p-value** |
| --- | --- | --- | --- | --- | --- |
| **Intercept** | 1.52 | 0.33 | 48.32 | 4.65 | <0.001* |
| **Pre-TMS Visit Bilateral Parietal** **DMN Connectivity** | -0.87 | 0.33 | 53.97 | -7.75 | <0.001* |
| **TMS Schedule** | -0.32 | 0.29 | 28.10 | -1.09 | 0.284 |
| **Group** | -0.38 | 0.37 | 44.52 | -1.01 | 0.318 |
| **TMS Schedule x Group** | 0.41 | 0.38 | 26.73 | 1.07 | 0.292 |

**Supplemental Table 6. Neural Effects of TMS Do Not Differ by TMS Administration Schedule*Group Interaction**. Model: Bilateral Parietal DMN Connectivity Change ~ Baseline Bilateral Parietal DMN Connectivity + group*schedule +(1|subid)

| **Predictors** | **Estimate** | **Std. Eror** | **df** | **t-value** | **p-value** |
| --- | --- | --- | --- | --- | --- |
| **Intercept** | 2.65 | 0.64 | 31.10 | 4.11 | <0.001* |
| **Pre-TMS Visit Bilateral Parietal** **DMN Connectivity** | -0.86 | 0.11 | 53.28 | -7.97 | <0.001* |
| **TMS Schedule** | -0.12 | 0.20 | 31.53 | -0.61 | 0.545 |
| **Group** | -0.48 | 0.33 | 27.68 | -1.43 | 0.163 |
| **Age** | -0.03 | 0.01 | 26.91 | -2.21 | 0.036* |

**Supplemental Table 7.** **Older age is associated with greater reductions in Bilateral Parietal** **DMN connectivity following DMN-targeted cTBS regardless of treatment schedule or diagnostic group.** Model: hyp1Aexpl = Bilateral Parietal DMN Connectivity Change ~ Baseline Bilateral Parietal DMN Connectivity + schedule + group + age + (1|subid)

| Adverse Event During or After TMS | Transdiagnostic Sample (n=34) | Number of Sessions of DMN-Targeted cTBS (total n=169^1^) |
| --- | --- | --- |
| Headache | 5 (14.71%) | 7 (4.14%) |
| Neck pain | 3 (8.82%) | 3 (1.78%) |
| Scalp pain/irritation | 1 (2.94%) | 4 (2.37%) |
| Hearing problems | 0 (0%) | 0 (0%) |
| Thinking problems | 0 (0%) | 1 (0.60%) |
| Concentration problems | 1 (2.9%) | 1 (0.60%) |
| Syncopal event or seizure | 0 (0%) | 0 (0%) |

**Supplemental Table 8. *Adverse Effects of Accelerated TMS Targeted to the Default Mode Network (DMN) in a Daily Schedule.*** After each session of TMS, participants were administered the TMS Acute Side Effects Questionnaire. Participants also completed the TMS Acute Side Effects one week after TMS during the follow-up MRI visit, and they also completed the questionnaire two weeks after TMS in a virtual follow-up. The results of the TMS Acute Side Effects Questionnaires administered directly following TMS are summarized in in the table above. There were no serious adverse events. Note^1^. One participant only completed 4 sessions of DMN TMS due to scheduling conflicts and not TMS side effects.

| Adverse Event During or After TMS | Transdiagnostic Sample (n=34) | Number of sessions of DMN-Targeted cTBS(total n=170) |
| --- | --- | --- |
| Headache | 4 (11.76%) | 5 (2.94%) |
| Neck pain | 0 (0%) | 0 (0%) |
| Scalp pain/irritation | 1 (2.94%) | 1 (0.59%) |
| Hearing problems | 0 (0%) | 0 (0%) |
| Thinking problems | 0 (0%) | 0 (0%) |
| Concentration Problems | 0 (0%) | 0 (0%) |
| Syncopal event or seizure | 0 (0%) | 0 (0%) |
| Other | 0 (0%) | 0 (0%) |
| Anxiety | 1 (2.94%) | 1 (0.59%) |
| Mood change | 1 (2.94%) | 2 (1.18%) |

**Supplemental Table 9. *Adverse Effects of Accelerated TMS Targeted to the Default Mode Network (DMN) in an Accelerated Schedule.*** After each session of TMS, participants were administered the TMS Acute Side Effects Questionnaire. Participants also completed the TMS Acute Side Effects one week after TMS during the follow-up MRI visit, and they also completed the questionnaire two weeks after TMS in a virtual follow-up. The results of the TMS Acute Side Effects Questionnaires administered directly following TMS are summarized in in the table above. There were no serious adverse events.
